# Is Fear of COVID-19 Higher among Food-Insecure Households? A Model-Based Study, Mediated by Perceived Stress among Iranian Populations

**DOI:** 10.1101/2020.12.22.20248714

**Authors:** Neda Ezzeddin, Hassan Eini-Zinab, Naser Kalantari, Mohammad Ahmadi, Zeinab Beheshti

**Author notes:** Corresponding Author Address: No. 7, Shahid Farahzadi Blvd, Shahid Hafezi St. (Western Arghavan), Ghods Town (West), Tehran,Iran., /.

## Abstract

The COVID-19 pandemic is a crisis accompanied with multiple psychological consequences. This cross-sectional study, conducted on 2871 Iranians, examines how food insecurity may be associated with fear of COVID-19. Household Food Insecurity Access Scale, COVID-19 fear scale, Cohen’s Perceived Stress Scale, and Perceived Social Support Questionnaire were used in gathering data. Data analysis was done using SPSS 22 and Amos 22. The results showed that food insecurity has significantly positive direct and indirect (mediated by perceived stress) correlations with fear of COVID-19 (P<0.05). It was also shown that perceived social support could negatively affect fear of COVID-19, through food insecurity or perceived stress (P<0.05). Among women, the presence of a child under 5 had a significant direct impact on fear of COVID-19 (P<0.05). The crisis caused by COVID-19 highlights the need for increasing social resilience through developing and implementing appropriate food security strategies.

**Statement of Relevance**
The COVID-19 pandemic is accompanied with multiple psychological consequences (including the *fear* of COVID-19), and threatens the food security status of millions of people. In order to assess the food insecurity impact on fear of COVID-19, the present study was conducted on Iranian adults. The proposed model, constructed by structural equation modeling, indicated the direct and indirect effects of food insecurity on fear of COVID-19. It was also shown that perceived social support could affect the fear of COVID-19 through food insecurity or perceived stress. It is concluded that food insecurity imposes further mental burdens during the pandemic, and policymakers should strengthen social resilience against its inter-related health consequences.

## Introduction

About a year ago, the first cases of COVID-19 were reported in Wuhan, China. This disease can cause death by chronic dysfunction of the lungs; it became a global pandemic due to high contagion rates (Salari et al., 2020). People’s mental health was also severely affected by the pandemic, for reasons such as the fear of getting infected, socio-financial disruptions, lockdowns, and so on (“The First COVID-19 Infanticide-Suicide Case,” 2020). Fear of COVID-19 is a mental health disorder (Pakpour & Griffiths, 2020), accompanied with different psychological consequences like anxiety, depression (Ahorsu et al., 2020), stress (Satici et al., 2020), and suicide in some severe cases (“Aggregated COVID-19 Suicide Incidences in India,” 2020; “The First COVID-19 Infanticide-Suicide Case,” 2020; Mamun & Griffiths, 2020). Fear of COVID-19 is also associated with child abuse (Tsur & Abu-Raiya, 2020) and lower life satisfaction (Satici et al., 2020).

The COVID-19 pandemic is a crisis which threatens the food security status of millions of people all over the world, due to its negative impacts on the global food system. The number of people with severe food insecurity is projected to nearly double due to the impact of COVID-19 by the end of the year 2020. Also, the number of malnourished children will increase due to increased wasting and stunting (FAO, 2020). Food insecurity has negative complications on health, including obesity and non-communicable diseases (NCDs) (Mosadeghrad et al., 2019), which in turn are associated with higher mortality and morbidity among COVID-19-infected people (Azarpazhooh et al., 2020; Kluge et al., 2020). It is also associated with other mental health conditions, including perceived stress (Martin et al., 2016). Perceived stress and fear of COVID-19 may lead to other serious mental health disorders, such as anxiety and depression (Rodríguez-Hidalgo et al., 2020). The purpose of the current study was to examine how food insecurity may be associated with fear of COVID-19 (a mental disorder which developed during the pandemic). This study was performed using structural equation modeling, mediated by perceived stress.

## Methods

### Study design, population and data gathering

This cross-sectional study was conducted on 2871 online Iranian people, from all 31 provinces nationwide; the proportion of geographical population distribution was considered. It ran from August to September 2020. The study population consisted of literate Iranian adults (18 years and older), who were selected by convenience sampling from popular social media platforms (namely Instagram, WhatsApp Messenger, and Telegram Messenger). All participants were included after approving an informed consent form.

## Measures

### Demographic and Socio-Economic Information

The demographic and socio-economic information of participants (including age, sex, household size, educational level, employment status, family residence status and household monthly income were obtained via online questionnaire. The participants were also asked about the presence of vulnerable people in the household. These included: pregnant woman, children under 5, elderly (over 65), and people with NCDs (such as cardiovascular disease, diabetes, cancer, endocrines disorder, etc.)

### Household Food Security Assessment

In this study, the Household Food Insecurity Access Scale (HFIAS) was used in order to determine the status of food insecurity among studied participants. This questionnaire consists of 9 questions with 4 frequencies of occurrences (most of the time; sometimes; rarely; and no), which are scored on a Likert scale (lowest score = zero, highest score = 3). The total scores obtained from the questionnaire are divided into 4 categories: food secure (0-1 score), mild food insecure (2-7), moderate food insecure (score 8-14) and severe food insecure (score 15-27). The validity and reliability of the Persian questionnaire has been previously assessed by Mohammadi et al. (Cronbach’s alpha=0.85) (Mohammadi et al., 2012).

### COVID-19 Fear Assessment

COVID-19 fear scale (FCV-19S) is a 7-item questionnaire that assesses the Fear of COVID-19 among participants, and is rated on a 5-point Likert scale (strongly disagree=1 to strongly agree=5). The total score is obtained by summing the scores of all individual items (score range 7-35); and the higher the rating, the more severe the fear. The reliability and validity of the Persian questionnaire was previously examined by Kwasi Ahorsu et al. (Cronbach’s alpha=0.82) (Ahorsu et al., 2020). An acceptable Cronbach’s alpha (0.87) was also obtained in the current study.

### Perceived Stress Assessment

The Cohen’s Perceived Stress Scale (PSS-14) is a self-report instrument was used to assess perceived stress among a study population. This questionnaire contains 14 items that examine the levels of thoughts and feelings of the person during the past month, and is scored on a 5-point Likert scale (Never=0; Almost Never=1; Sometimes=2; Fairly Often=3; Very Often=4). Questions (4-7), (9-10) and 13 are scored in reverse. In this scale, the minimum perceived stress score is (0) and the maximum is (56). The higher the score, the more the perceived stress (L. Mohammadi-Yeganeh et al., 2008). Acceptable internal reliability for the current questionnaire was previously obtained by Qazvini et al. (Cronbach’s alpha=0.73) (Ghasedi Qazvini & Kiani, 2018).

### Perceived Social Support Assessment

In the current study, the Multidimensional Scale of Perceived Social Support Questionnaire (MSPSS) was used in order to assess the perceived social support. This 12-item scale consists of three subscales, examining the perceived social support from three sources (family, friends, and others), and is scored on a 5-point Likert scale (strongly disagree=1 to strongly agree=5). The total score is obtained by summing all the individual item scores. The minimum and maximum perceived social support score is (12) and (60), respectively. Cronbach’s alpha of 0.93 was obtained for the Persian MSPSS questionnaire by Salimi et al. (A.r et al., 2009).

### Statistical Analysis

Descriptive and analytical analysis (including: Pearson correlation, T-test, ANOVA test and linear regression) were performed using IBM SPSS Statistics for Windows, Version 22.0; and IBM SPSS Amos Version 22.0 for structural equation modeling (SES).

## Results

The mean age of participants was 32.99±8.31, and many of them were employees (24.7%), with associate or bachelor’s degree (47.1%). The detailed characteristics of the participants are presented in Table 1. Since the frequency of women in the raw data was much higher than men (82.8% vs. 17.2%), data weighting was done by sex, according to the latest census of the Statistics Center of Iran (51% men, 49% women).

**Table 1.**
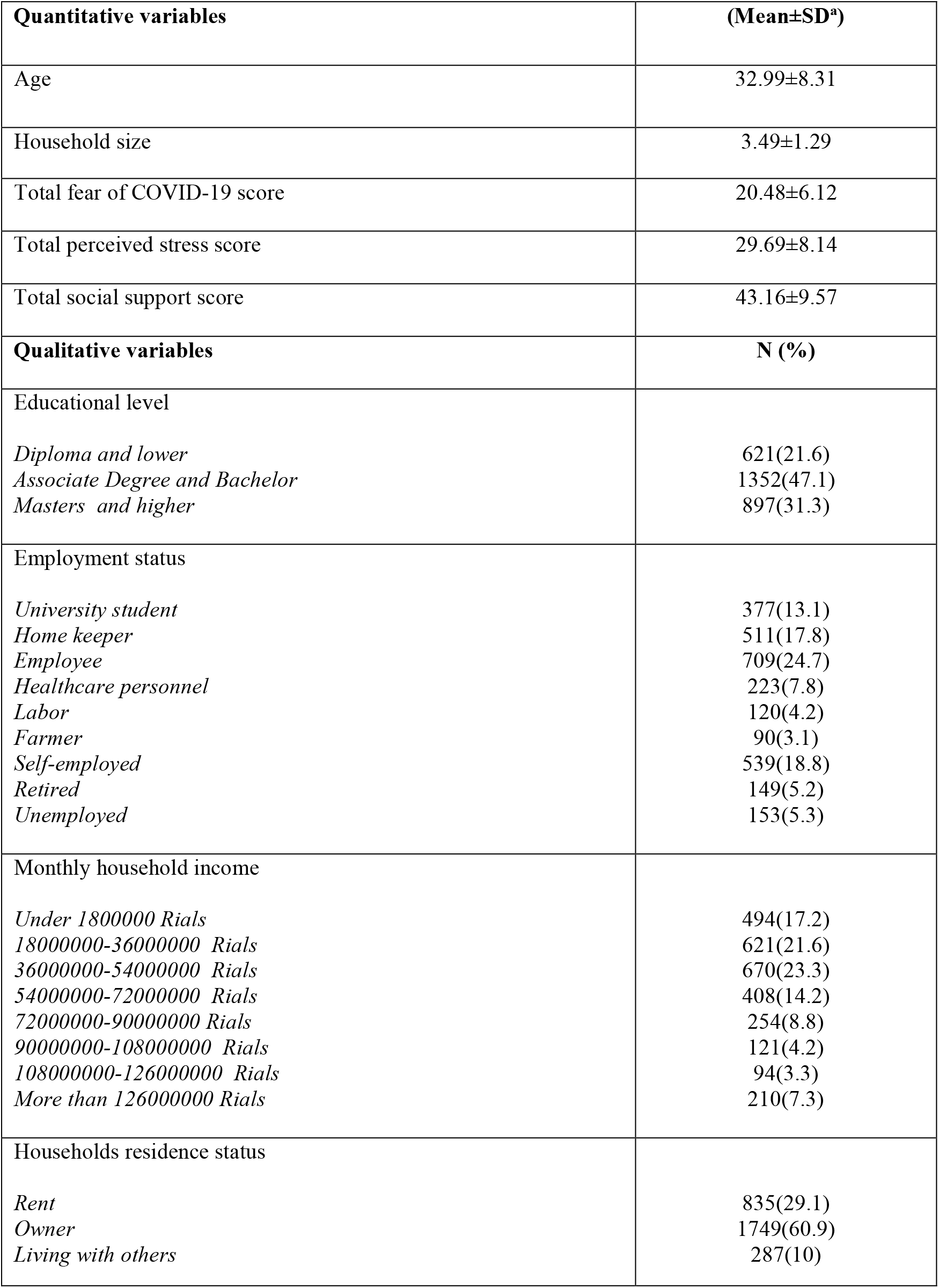
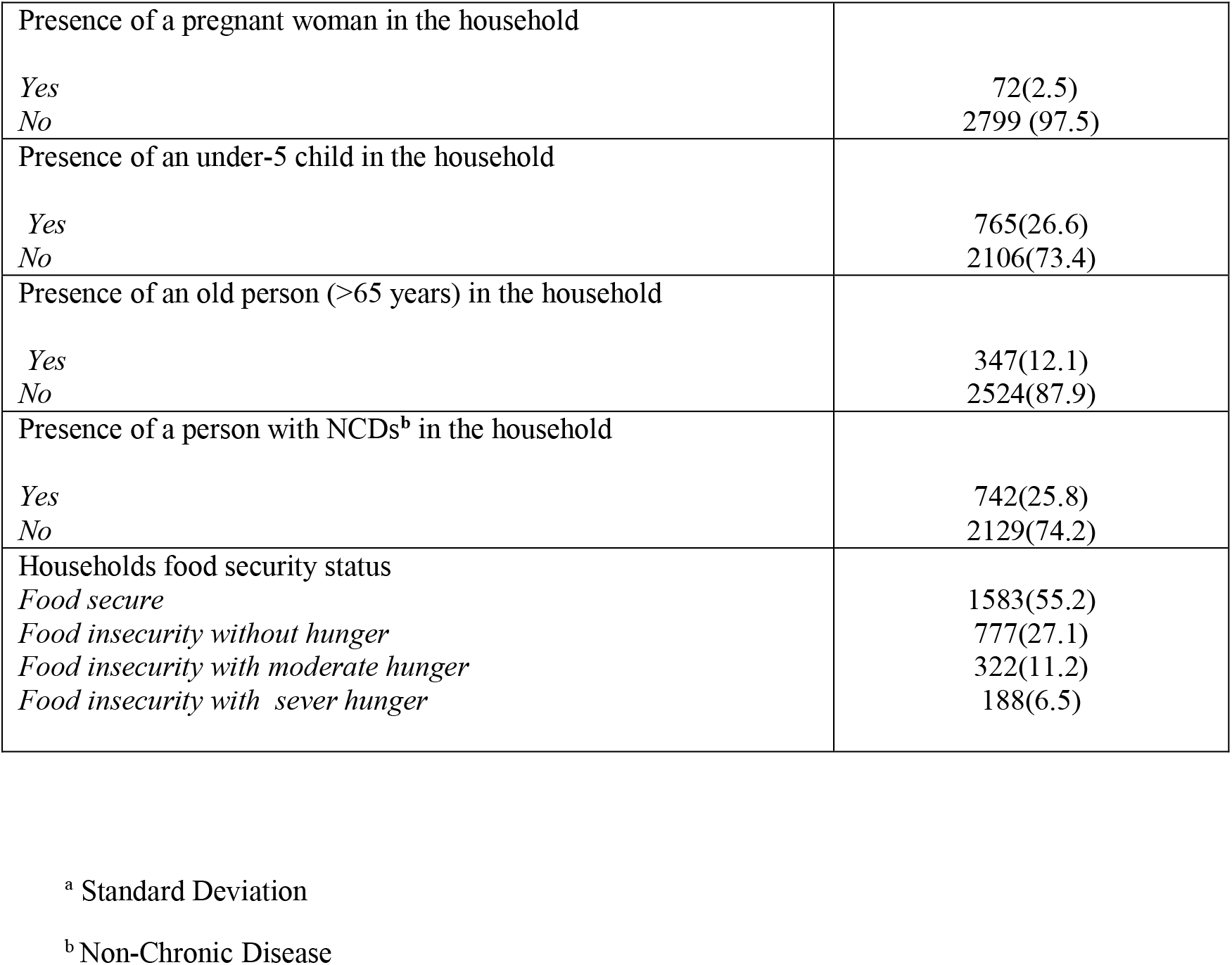
The characteristics of the participants

| Quantitative variables | (Mean±SD<sup>a</sup>) |
| --- | --- |
| Age | 32.99±8.31 |
| Household size | 3.49±1.29 |
| Total fear of COVID-19 score | 20.48±6.12 |
| Total perceived stress score | 29.69±8.14 |
| Total social support score | 43.16±9.57 |
| Qualitative variables | N (%) |
| Educational level |  |
| *Diploma and lower* | 621(21.6) |
| *Associate Degree and Bachelor* | 1352(47.1) |
| *Masters and higher* | 897(31.3) |
| Employment status |  |
| *University student* | 377(13.1) |
| *Home keeper* | 511(17.8) |
| *Employee* | 709(24.7) |
| *Healthcare personnel* | 223(7.8) |
| *Labor* | 120(4.2) |
| *Farmer* | 90(3.1) |
| *Self-employed* | 539(18.8) |
| *Retired* | 149(5.2) |
| *Unemployed* | 153(5.3) |
| Monthly household income |  |
| *Under 1800000 Rials* | 494(17.2) |
| *18000000-36000000 Rials* | 621(21.6) |
| *36000000-54000000 Rials* | 670(23.3) |
| *54000000-72000000 Rials* | 408(14.2) |
| *72000000-90000000 Rials* | 254(8.8) |
| *90000000-108000000 Rials* | 121(4.2) |
| *108000000-126000000 Rials* | 94(3.3) |
| *More than 126000000 Rials* | 210(7.3) |
| Households residence status |  |
| *Rent* | 835(29.1) |
| *Owner* | 1749(60.9) |
| *Living with others* | 287(10) |
| Presence of a pregnant woman in the household |  |
| *Yes* | 72(2.5) |
| *No* | 2799 (97.5) |
| Presence of an under-5 child in the household |  |
| *Yes* | 765(26.6) |
| *No* | 2106(73.4) |
| Presence of an old person (>65 years) in the household |  |
| *Yes* | 347(12.1) |
| *No* | 2524(87.9) |
| Presence of a person with NCDs<sup>b</sup> in the household |  |
| *Yes* | 742(25.8) |
| *No* | 2129(74.2) |
| Households food security status |  |
| *Food secure* | 1583(55.2) |
| *Food insecurity without hunger* | 777(27.1) |
| *Food insecurity with moderate hunger* | 322(11.2) |
| *Food insecurity with sever hunger* | 188(6.5) |

<sup>a</sup> Standard Deviation
<sup>b</sup> Non-Chronic Disease

In the current study, socio-economic variables (including household residence status, monthly income, participant’s educational level, and employment status) were analyzed by the Principal Component Analysis (PCA) method [Kaiser-Meyer-Olkin test (KMO)>0.5 and Bartlett test<0.001]. A socio-economic status variable was developed and split into three levels. Based on the results, more than half of the participants (55.2%) were from food-secure households, and about 6.5% were reported as severely food-insecure. For subsequent analyses, the third and fourth groups (food-insecure with moderate and severe hunger) were merged.

The Pearson correlation showed a significant association between fear of COVID-19 with quantitative variables, including age (r=0.05, P<0.05), total perceived stress score (r=0.37, P<0.001), and total social support score (r=-0.05, P<0.05). However, household size was not an important predictor of fear of COVID-19 (r=0.01, P>0.05). The analysis by T-test showed that the mean fear of COVID-19 score was significantly higher among women (P<0.001); in households with a child under 5; and in patients with NCDs (P<0.05) (Table 2). The association of fear of COVID-19 score and household food security status indicated that with increasing degree of food insecurity, the fear of COVID-19 score also increases (P<0.001). However, the ANOVA test did not show any significant association between fear of COVID-19 and the level of socio-economic status (P>0.05) (Table 2).

**Table 2.** The association of fear of COVID-19 with qualitative variables

| Variables | Total fear of COVID-19 score (Mean ±SD<sup>a</sup>) | 95% CI<sup>b</sup> | P-value |
| --- | --- | --- | --- |
| Sex |  |  |  |
| Women | 21.67±5.93 | 1.89, 2.77 | <0.001* |
| Men | 19.33±6.08 | | |
| Presence of a pregnant woman in the household |  |  |  |
| Yes | 20.14±6.28 | -1.09, (1.78) | >0.05* |
| No | 20.49±6.12 | | |
| Presence of an under-5 child in the household |  |  |  |
| Yes | 21.10±6.48 | -1.37, (-0.32) | <0.05* |
| No | 20.25±5.97 | | |
| Presence of an old person (>65 years) in the household |  |  |  |
| Yes | 20.94±5.98 | -1.21, (0.15) | >0.05* |
| No | 20.41±6.14 | | |
| Presence of a person with NCDs<sup>c</sup> in the household |  |  |  |
| Yes | 20.92±6.06 | -1.10, (-0.07) | <0.05* |
| No | 20.33±6.14 | | |
| Households food security status |  |  |  |
| Food secure | 19.60±5.95 | 20.25, 20.70 | <0.001** |
| Food insecurity without hunger | 21.28±6.05 | | |
| Food insecurity with moderate- sever hunger | 21.99±6.30 | | |
| Scio-economic status |  |  |  |
| First tertile | 20.49±6.06 | 20.25, 20.70 | >0.05** |
| Second tertile | 20.74±6.15 | | |
| Third tertile | 20.21±6.14 | | |

\* T-test
\*\* ANOVA
<sup>a</sup> Standard Deviation
<sup>b</sup> Confidence Interval
<sup>c</sup> Non-Chronic Disease

In the next step, a linear regression model was used for predicting fear of COVID-19, by including the significant variables (P<0.05) identified by Pearson correlation, T-test and ANOVA test. The results showed that age, sex, perceived social support, perceived stress, food security status and the presence of a child under 5 were significant predictors of fear of COVID-19 (P<0.05) (Table 3).

**Table 3.** Final model of linear regression to examine the association between the fear of COVID-19 and household food security status

| Variable | B | SE <sup>a</sup> | $\beta$ | t | 95% CI<sup>b</sup> | P- value |
| --- | --- | --- | --- | --- | --- | --- |
| age | 0.80 | 0.013 | 0.109 | 6.381 | 0.055, 0.105 | <0.001 |
| Total social support score | 0.058 | 0.012 | 0.091 | 4.857 | 0.035, 0.81 | <0.001 |
| Total perceived stress score | 0.278 | 0.014 | 0.370 | 19.352 | 0.250, 0.306 | <0.001 |
| Household Food security status | 0.757 | 0.146 | 0.095 | 5.199 | 0.471, 1.042 | <0.001 |
| Presence of a person with NCDs<sup>c</sup> in the household | 0.243 | 0.241 | 0.017 | 1.007 | -0.230, 0.716 | >0.05 |
| Presence of an under-5 child in the household | 0.635 | 0.239 | 0.046 | 2.656 | 0.166, 1.104 | <0.05 |
| sex | -1.582 | 0.214 | -0.129 | -7.377 | -2.002, (-1.161) | <0.001 |
| Notice: R= 0.429 R<sup>2</sup>= 0.184 ADJ.R<sup>2</sup><sup>d</sup>= 0.182 |  |  |  |  |  |  |

<sup>a</sup> standard error
<sup>b</sup> Confidence Interval
<sup>c</sup> Non-Chronic Disease
<sup>d</sup> Adjusted R Square

Finally, the structural equation modeling (SES) of fear of COVID-19 was constructed using the significant variables. Figures 1 and 2 show the pathways by groups (sex), for men and women respectively. The proposed model (by sex) includes three exogenesis variables (age, perceived social support and the presence of a child under 5), hierarchical mediator variables (food security status and perceived stress), and an endogenous variable (fear of COVID-19). The results of the model indicated that all pathways in both sexes are significant, except for having a child under 5, which was significant only in the female group (Table 4). There was also a significant sex difference between fear of COVID-19 and food security status pathways (P<0.05), as it was seen to be stronger among men. A comparison of the between-pathways coefficient by sex also showed a greater inverse association between age and fear of COVID-19 among men. The standardized total, direct and indirect effects are provided in Table 4, and confirm the indirect effect of food insecurity on fear of COVID-19, in terms of perceived stress. Acceptable fit indices were obtained for the model, which are presented in Table 4 (end of table).

**Table 4.** Summary of the model for predicting the fear of COVID-19, by sexes

| **Women group** | **Standardized Direct Effects** | **Standardized Indirect Effects** | **Standardized Total Effects** | **Estimate** | **S.E.** | **C.R.** | **P-value** |
| --- | --- | --- | --- | --- | --- | --- | --- |
| **pathways** |  |  |  |  |  |  |  |
| Fear of COVID-19 <---age | 0.090 | -0.054 | 0.036 | 0.067 | 0.018 | 3.619 | <0.001 |
| Fear of COVID-19 <--- perceived social support | 0.084 | -0.167 | -0.083 | 0.052 | 0.017 | 3.134 | 0.002 |
| Fear of COVID-19 <--- having under-5 child | 0.085 | 0.000 | 0.085 | 1.129 | 0.327 | 3.451 | <0.001 |
| Perceived stress <--- Food security status | 0.120 | 0.000 | 0.120 | 1.384 | 0.293 | 4.719 | <0.001 |
| Fear of COVID-19 <--- Food security status | 0.095 | 0.045 | 0.140 | 0.775 | 0.145 | 5.329 | <0.001 |
| Fear of COVID-19 <--- Perceived stress | 0.375 | 0.000 | 0.375 | 0.268 | 0.019 | 13.991 | <0.001 |
| **Men group** | **Standardized Direct Effects** | **Standardized Indirect Effects** | **Standardized Total Effects** | **Estimate** | **S.E.** | **C.R.** | **P-value** |
| **pathways** |  |  |  |  |  |  |  |
| Fear of COVID-19 <---age | 0.131 | -0.030 | 0.101 | 0.092 | 0.017 | 5.433 | <0.001 |
| Fear of COVID-19 <--- perceived social support | 0.102 | -0.167 | -0.065 | 0.066 | 0.017 | 3.927 | <0.001 |
| Fear of COVID-19 <--- having under-5 child | 0.003 | 0.000 | 0.003 | 0.039 | 0.333 | 0.117 | 0.907 |
| Perceived stress <--- Food security status | 0.294 | 0.000 | 0.294 | 2.818 | 0.229 | 12.281 | <0.001 |
| Fear of COVID-19 <--- Food security status | 0.102 | 0.108 | 0.210 | 0.775 | 0.145 | 5.329 | <0.001 |
| Fear of COVID-19 <--- Perceived stress | 0.367 | 0.000 | 0.367 | 0.291 | 0.021 | 13.750 | <0.001 |
| *fit indices* | **TLI** | **GFI** | **AGFI** | **IFI** | **CFI** | **RMSEA** |  |
|  | 0.93 | 0.99 | 0.98 | 0.97 | 0.97 | 0.03 |  |

TLI=Tucker-Lewis index; GFI =Goodness of Fit Index; AGFI = adjusted goodness of fit index; IFI = Incremental Fit Index; CFI=comparable fit index; RMSEA=Root Mean Square Error of Approximation

**Figure 1:**
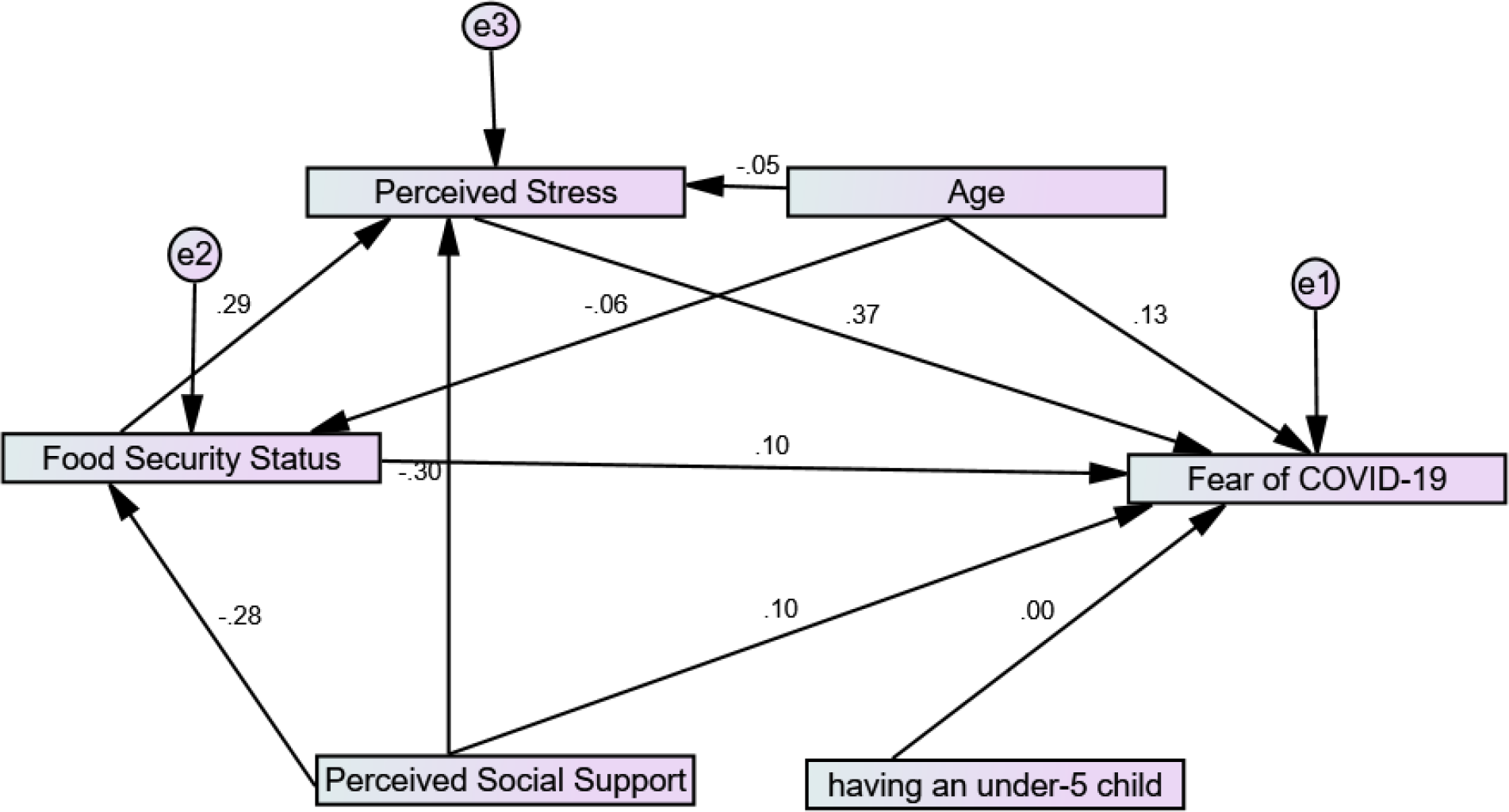
proposed model for predicting the fear of COVID-19, in men

**Figure 2:**
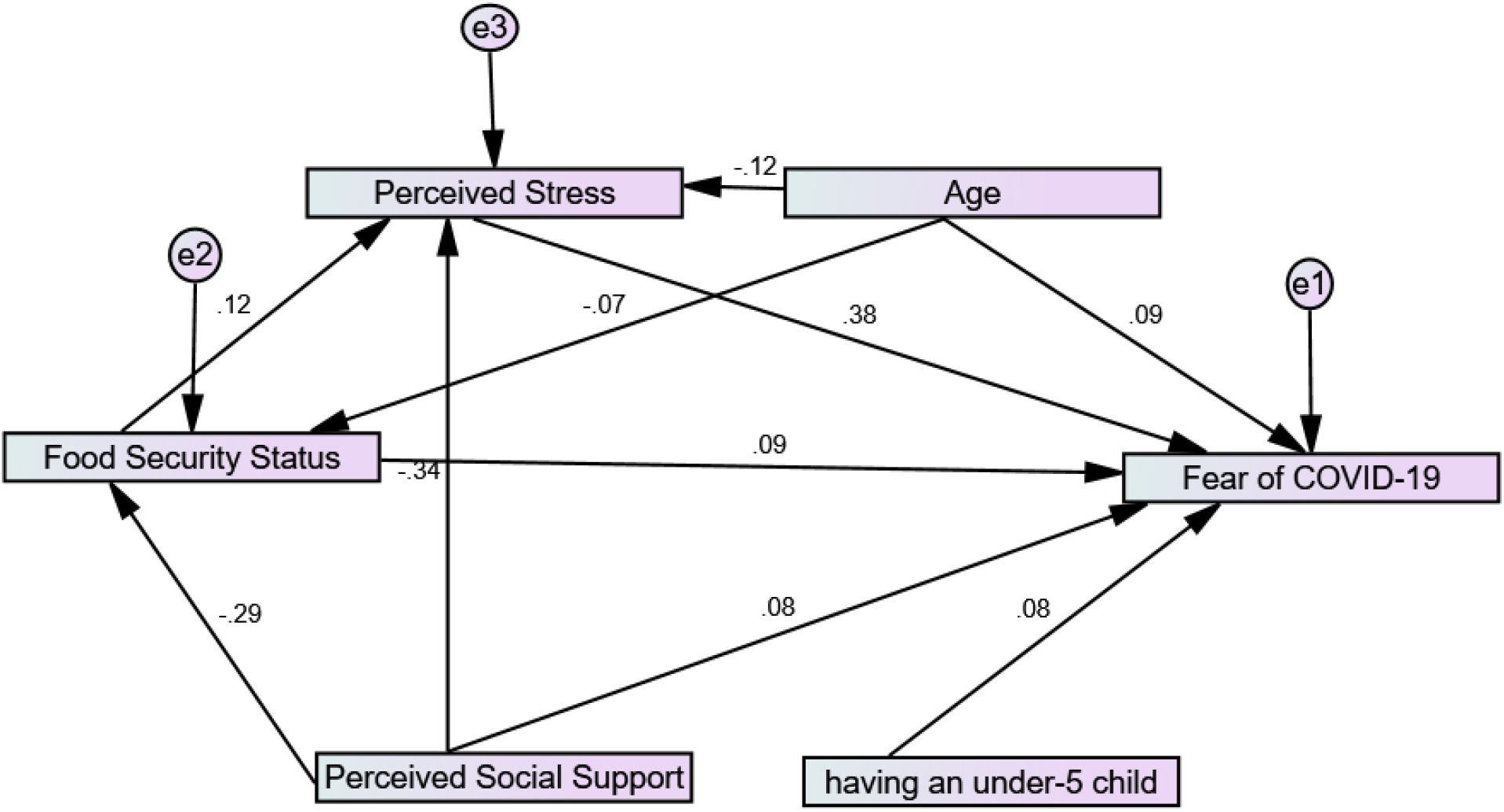
proposed model for predicting the fear of COVID-19, in women

## Discussion

The current study was conducted among an Iranian online population, in order to determine the impact of food insecurity on fear of COVID-19, mediated by perceived stress in structural equation modeling. The results indicated the presence of both direct and indirect effects of food insecurity on fear of COVID-19. It was also shown that perceived social support could affect fear of COVID-19, through food insecurity or perceived stress.

Investigating the relationship between food insecurity and mental health disorders is an ongoing topic of interest to researchers. It has been shown that depressive symptoms, anxiety and stress are higher among food-insecure households (Ezzeddin et al., 2018; Tribble et al., 2020). Food-insecure households usually consume low quality diets, which in turn, are associated with poor mental health (Davison et al., 2017). Worries about family food sources can also be stressful (Hromi-Fiedler et al., 2009). In the current study, food insecurity was significantly associated with higher perceived stress and fear of COVID-19. The COVID-19 pandemic has created conditions that put additional stress on people (Alkhamees et al., 2020). Taylor et al. showed that the concerns about meeting household needs, due to the social and economic disruption following the COVID-19 outbreak, can be associated with perceived stress (Taylor et al., 2020). In another study conducted by Rehman et al. among Indians during COVID-19 pandemic, it was shown that an insufficient food supply during quarantine was associated with greater mental distress, including perceived stress (Rehman et al., 2020). The COVID-19 pandemic has left many people with job losses, reduced income, (Hirvonen et al., 2020) and increased food prices (FAO, 2020), raising concerns about the growing prevalence of food insecurity. It has been shown that these economic problems in households are associated with insufficient food intake, both in terms of amount and quality. However, families with persistent income or enough savings were seen to be less affected during the quarantine period (Ruszczyk et al., 2020). Food insecurity also makes people physically (Kelly et al., 2018) and mentally (Slavich, 2020) more vulnerable. In a study conducted by Kelly et al., the death risk from Ebola was 18.3 times higher among food-insecure patients (Kelly et al., 2018). A sufficient and nutritious diet plays an important role in stimulating an appropriate immune system response to COVID-19 (Iddir et al., 2020). Stress reduction, nutritious diet, adequate levels of vitamin D in the blood, and adequate physical activity are other important factors associated with better immune system function (Wimalawansa, 2020). Long-term fear and stress due to COVID-19 can also alter the body’s neuro-endocrine-immune system, which may lead to an increase in other diseases (Liu et al., 2020). In order to reduce the effects of food insecurity on various aspects of health, then, policymakers and planners need to develop both short- and long-term strategies to increase community resilience to such shocks. Inevitably, this requires cooperation and partnership at the global, national, and local levels.

According to the results of the current study, perceived social support affects fear of COVID-19 via different pathways: direct and indirect. The indirect effect of social support was such that, as the score increased, perceived stress decreased. This finding is consistent with a study conducted by Ye et al. among college students in China (Ye et al., n.d.). It has been shown that social support is also an important factor negatively associated with perceived stress and anxiety among healthcare workers during the pandemic (Labrague & Santos, 2020; Xiao et al., 2020). Social support has played an influential role on mental health and wellbeing during the pandemic, but unfortunately, it has suffered due to social distancing practices (El-Zoghby et al., 2020; Saltzman et al., 2020). Access to technology (and its functions in maintaining social relationships) may be able to mitigate feelings of loneliness and mental health problems, however (Saltzman et al., 2020). Another indirect pathway that links perceived social support to fear of COVID-19 is through its positive impact on food security. This finding is consistent with studies by Ashe (Ashe & Lapane, 2018) and Nascimento Dos Santos (Santos et al., 2019). People with higher social support are more likely to be wealthy, and less likely to experience food insecurity (Hadley et al., 2007).

### Limitations

This cross-sectional study was conducted among an Iranian online population, through popular social networks. Currently, no previous study has examined the relationship between household food insecurity and fear of COVID-19 (or the research is limited), so the present study is innovative in terms of subject matter. Another strength of this study was its use of different variables, and the assessment of their interaction within the model. However, the study had some limitations, including the lower participation of older people (over 65). Convenience sampling, which was used in data gathering, may be accompanied with bias and lower generalizability (Ahorsu et al., 2020).

In order to address these limitations, this study utilized a relatively high sample size, a proportional approach to geographical distribution, and data weighting.

## Conclusion

In addition to threatening people’s physical health, the pandemic threatens food security and mental health. Based on the results of the present study, food insecurity has significant direct and indirect (mediated by perceived stress) impacts on fear of COVID-19. The crisis caused by COVID-19 highlights the need to increase social resilience through developing and implementing appropriate strategies to prevent and mitigate social costs, whether they be physical, psychological, or nutritional. This study showed that increasing food security resilience can play a key role in achieving these goals.

## Data Availability

According to the research project contract, the researchers are not allowed to share the data directly, but the data will be available through correspondence with the Vice-Chancellor of Research Affairs

## Acknowledgments

The authors would like to thank the Vice-Chancellor of Research Affairs, Shahid Beheshti University of Medical Sciences, for funding assistance (code: 413, date of approval: 2 August 2020). The authors are thankful to the participants in the study, and those who shared the questionnaire link via social media.

## Author Contributions

NE participated in research design, gathering and analyzing the data, and preparing the manuscript. HE participated in data analysis and critically reviewing the manuscript. NK participated in research design and critically reviewing the manuscript. MH and ZB participated in data gathering. The manuscript content was approved by all authors.

## Ethics Approval

This study was approved by the Ethics Committee of the National Nutrition and Food Technology Research Institute, Shahid Beheshti University of Medical Sciences, 26 July 2020 (ethical code: IR.SBMU.nnftri.Rec.1399 .028).

